# Timing and magnitude of the next wave of COVID-19 in China: lessons from 189 countries and territories

**DOI:** 10.1101/2023.03.27.23287793

**Authors:** Beidi Niu, Shuyi Ji, Shi Zhao, Hao Lei

**Affiliations:** School of Public Health, Zhejiang University, Hangzhou 310058, China; JC School of Public Health and Primary Care, Chinese University of Hong Kong, Hong Kong SAR 999077, China; The Key Laboratory of Intelligent Preventive Medicine of Zhejiang Province, Hangzhou, Zhejiang 310058, China

## Abstract

Because of the fading immunity to COVID-19 and continuous evolution of the SARS-CoV-2 Omicron variants, the next epidemic wave of COVID-19 is inevitable. The Omicron variant has been the cause of several waves of the COVID-19 epidemics in the majority of countries. Thus, lessons from other countries may provide guidance regarding the timing and magnitude of the next COVID-19 wave of the pandemic in China. In this study, the COVID-19 surveillance data from 189 countries that experienced two or more waves of the SARS-CoV-2 Omicron variant were analysed. The median peak timing between the first and second/third waves of the SARS-CoV-2 Omicron variant was 164/243 days. The peaks of the second and third waves were much lower than that of the first wave. The median relative peaks of the second and third compared with the first waves were 14.5% and 11.2%, respectively. The time window between the peak timings of the first and second waves showed no significant rank correlation with the five socioeconomic factors included in this study. However, the relative peak of the second wave increased significantly with gross domestic product per capita (*P*<0.001), urbanisation rate (*P*=0.003), population density (*P*=0.007), and proportion of older adults >65 years (*P*<0.001), although decreased significantly with the proportion of 0-14 teenagers (*P*<0.001). In summary, the historical situations and progression of COVID-19 outbreaks in other countries may inform the risk assessment of incoming outbreaks in mainland China; however, the timing and magnitude of the next COVID-19 wave may also be influenced by several unknown factors, including rapid viral evaluation of SARS-CoV-2

## Background

Knowledge of the timing and magnitude of the next wave of the coronavirus disease 2019 (COVID-19) pandemic in China is important for predicting and preventing the spread of infection. The most serious was also the first nationwide wave of COVID-19, and was mainly caused by the severe acute respiratory syndrome coronavirus 2 (SARS-CoV-2) Omicron BF.7 and BA.5.2 variants, which had nearly disappeared by 25 January 2023 in China [0]. However, because the immunity to COVID-19 fades [1] and the SARS-CoV-2 Omicron variant continues to evolve [2], the next COVID-19 wave will happen sooner or later. The human body learns to recognize SARS-CoV-2 through vaccination, infection, or both. However, existing findings in the literature suggested that the immunity against COVID-19 would wane with a relatively high decay rate, regardless of previous vaccine or infection history [1]. A review suggested that hybrid immunity, that is, immunity acquired through both vaccination and infection, provides longer immunity, although the effectiveness of hybrid immunity still waned to 41.8% (95% confidence interval [CI] 31.5–52.8) after 12 months [4]. The effectiveness of previous infection against reinfection was much lower, and waned to 24.7% (95% CI 16.4–35.5) after 12 months [4].

Mainly owing to the zero-COVID policy and strict nonpharmaceutical interventions implemented in China, the first nationwide COVID-19 wave caused by the Omicron variant occurred in China much later compared with other countries. Since the Omicron variant was first identified in South Africa and Botswana and was reported to the World Health Organization (WHO) on 24 November 2021 [5], it has caused at least three COVID-19 waves in Europe, the Americas, and the West Pacific [6]. Thus, lessons from other countries may provide clues regarding the timing and magnitude of the next COVID-19 wave in China.

In this study, we analysed COVID-19 surveillance data from 237 countries to quantify the timing and magnitude of the first and second waves of COVID-19 caused by the SARS-CoV-2 Omicron variant. In addition, we explored the correlations with socioeconomic factors and predicted the timing and magnitude of the next COVID-19 wave in China.

## Methods

### Data sources

The number of daily confirmed COVID-19 cases from 237 counties between 1 January 2020 and 28 February 2023 was extracted from the WHO Coronavirus (COVID-19) Dashboard [6], Johns Hopkins University Center for Systems Science and Engineering [7], and Our World in Data [8]. Moreover, we collected socioeconomic status data for each country from The World Bank [5], including the gross domestic product (GDP) per capita, urbanisation rate (percentage of people living in urban areas), population density, and the proportions of people aged >65 years and <14 years.

### Data analysis

Since the SARS-CoV-2 Omicron variant was first identified in South Africa and Botswana and was reported to the WHO on 24 November 2021 [9], it has spread rapidly and become and remained the dominant variant worldwide to date. Therefore, in this study, we focused on the period after 24 November 2021.

In this study, the period between two consecutive waves of the SARS-CoV-2 Omicron variant was defined as the period between the peak timing of these two consecutive waves. The daily peak number of confirmed COVID-19 cases in each wave was recorded. The relative peak of the second, third, and fourth waves was defined as the peak of the second, third, and fourth waves divided by the peak of the first wave. Since the first nationwide wave of the SARS-CoV-2 Omicron variant occurred in January 2023 in China, the timing of the second wave of the SARS-CoV-2 Omicron variant is of great concern. Thus, we mainly focused on the period between the first and second waves of the SARS-CoV-2 Omicron variant in other countries. The definition of a COVID-19 wave within a country used in this study was similar to previous studies on influenza [10, 11], the onset timing of an epidemic wave was defined as the first seven consecutive days with a smoothed increase in the number of daily cases exceeding a prescribed baseline. The baseline was set as the 50% quantile of the non-zero number of daily cases. The end of a wave was defined as the first of seven consecutive days with a smoothed number of daily cases below the baseline. Because fluctuations in surveillance data may play a particular role in estimating the peak timing and peak value of a wave, a 7-day moving average was used to smooth the COVID-19 surveillance data in this study [10].

Spearman’s correlation was used to characterise the correlation between socioeconomic factors and the period of peak timing between two consecutive waves, or the relative peak of the second wave, as these data did not follow a normal distribution.

### Modelling study

However, Even until now, the predictingprediction of the transmission dynamics of the SARS-CoV-2 remains challengingSARS-CoV-2 is still difficult. Therefore, So in this study, we built a simple susceptible-exposed-infected-recovered-susceptible (SEIRS) model to explore the relationship between immunity periods and periods between the peak timing of the first and second waves of the SARS-CoV-2 Omicron variant. The SEIRS model can be represented by a set of ordinary differential equations as follows:

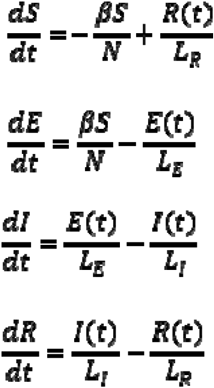

Where ***S(t), E(t)***, ***I(t)***, and ***R(t)*** denoted the number of susceptible, exposed, infected, and recovered individuals at time *t*, respectively. ***N*** is the total population size, and ***N =*** 1412600000 [12]; ***L***_***E***_, ***L***_***I***_ and ***L***_***R***_ represent the latent, infectious, and immunity periods, respectively;. ***L***_***E***_ = 3.8 days [13], ***L***_***I***_ = 5 days [14]. ***β*** is the transmission rate of SARS-CoV-2 transmission rate, and was estimated by the least-squares method based on the number of daily COVID-19 cases in China between 16 November 2022 and 14 February 2023 in accordance with the WHO [6]. According to the WHO, during this period, the total number of COVID-19 cases was 89.46 million, accounting for 6.3% of the total population in China. However, based on seroprevalence data in Guangzhou, in China as of 14 January, 2023 the overall attack rate of SARS-CoV-2 was estimated to be 61.5% [15], which is approximately 10 times the rate of 6.3% reported by the WHO. Thus, we simply expanded the daily number of COVID-19 cases reported by the WHO by 10 and smoothed the data by a 7-day moving average.The reported immunity period against SARS-CoV-2 varies greatly in different studies, ranging from 6 to > 20 months [16-19]. We performed a sensitivity analysis and selected three values: 6, 12, and 20 months.

## Results

A total of 237 counties were included in this study. Based on our definition of a COVID-19 wave, by March 2023, two or more waves of the SARS-CoV-2 Omicron variant had occurred in 189 countries, while three or more waves occurred in 120 countries, and in 48 countries, more than four waves had happened (Figure 1). In the 189 countries with two or more waves of COVID-19, the detailed timing and peak number of daily cases in each wave of COVID-19 caused by the SARS-CoV-2 Omicron variant, socioeconomic status, and COVID-19 vaccination rate are listed in the Supplementary Material.

**Figure 1.**
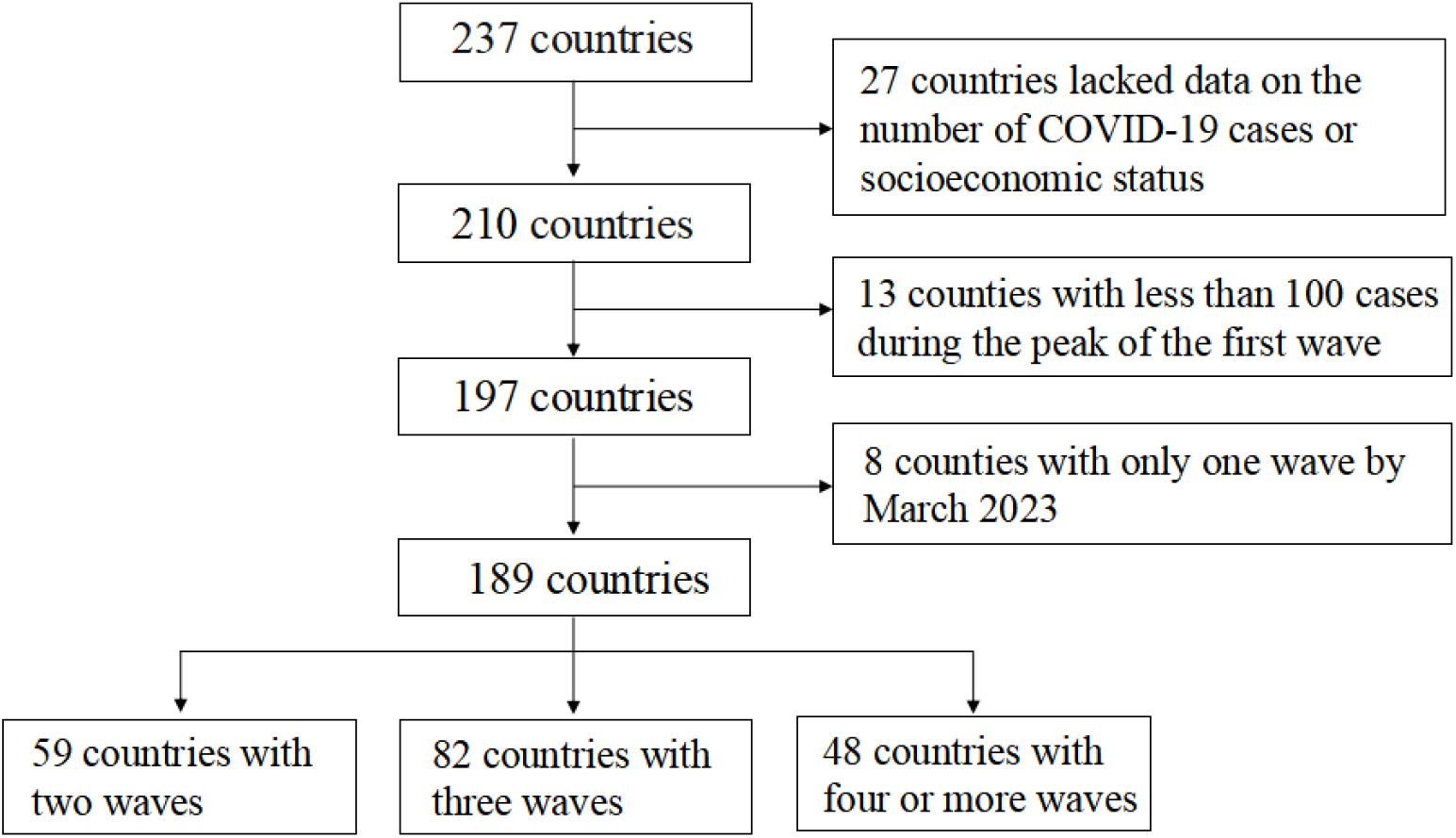
The selection process of countries included in this study.

The peak timings of the first and second waves varied significantly across 189 countries (Figure 2). The SARS-CoV-2 Omicron variant was first identified in South Africa, and the peak timing of the first wave occurred early, on 15 December 2021. The early peak timing of the first wave did not always indicate early peak timing of the second wave (Figure 2).

**Figure 2.**
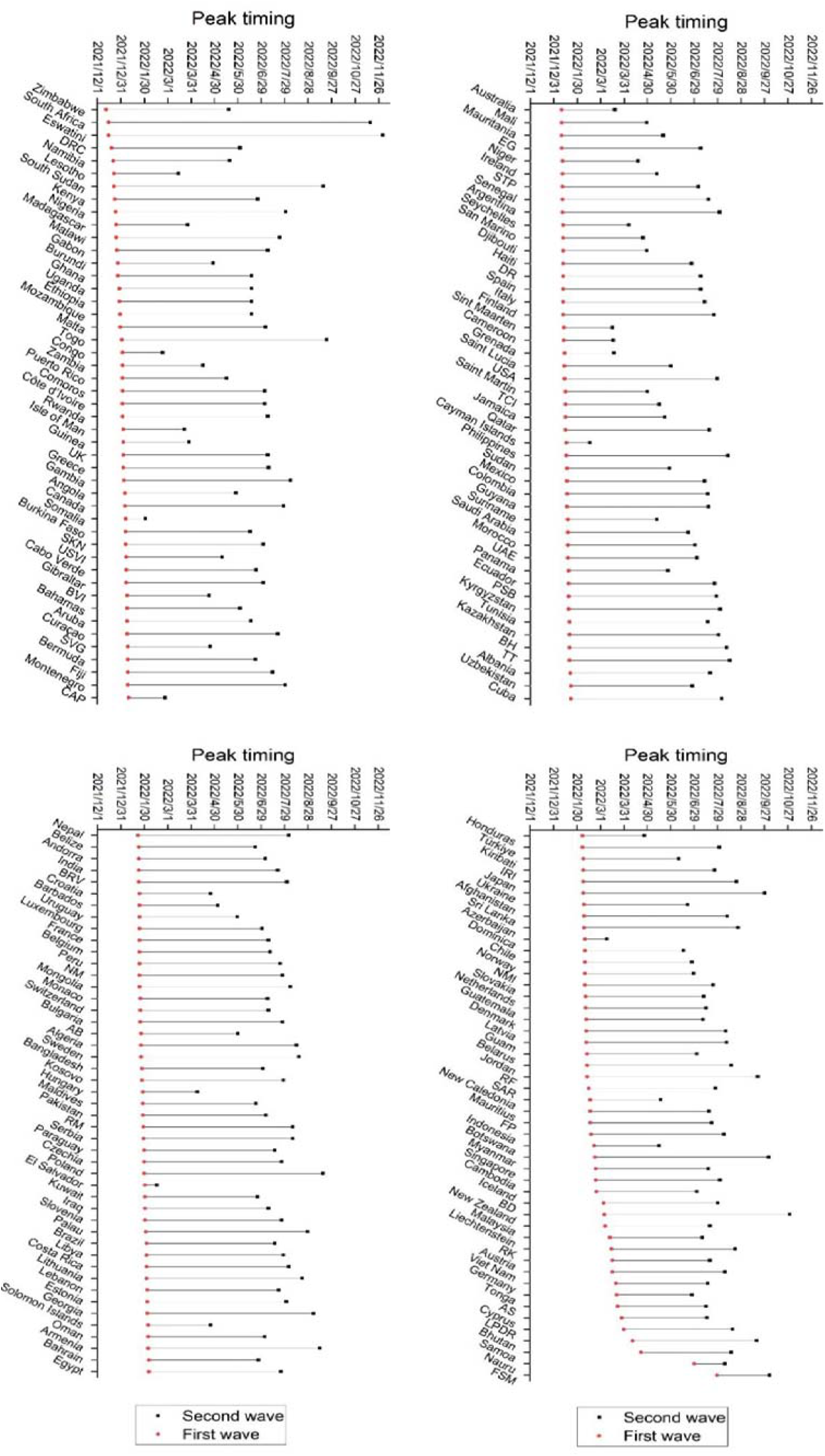
Peak timing of the first and second waves of COVID-19 caused by the SARS-CoV-2 Omicron variant in 189 countries. Countries were sorted by the increased peak timing of the first waves. Abbreviations: Democratic Republic of the Congo, DRC; The United Kingdom, UK; Saint Kitts and Nevis, SKN; United States Virgin Islands, USVI; British Virgin Islands, BVI; Saint Vincent and the Grenadines, SVG; Central African Republic, CAP; Equatorial Guinea, EG; Sao Tome and Principe, STP; Dominican Republic, DR; United States of America, USA; Turks and Caicos Islands, TCI; United Arab Emirates, UAE; Bolivia (Plurinational State of), PSB; Bosnia and Herzegovina, BH; Trinidad and Tobago, TT; Venezuela (Bolivarian Republic of), BRV; North Macedonia, NM; Antigua and Barbuda, AB; Republic of Moldova, RM; Iran (Islamic Republic of), IRI; Northern Mariana Islands (Commonwealth of the), NMI; Russian Federation, RF; Syrian Arab Republic, SAR; French Polynesia, FP; Brunei Darussalam, BD; Republic of Korea, RK; American Samoa, AS; Lao People’s Democratic Republic, LPDR; Micronesia (Federated States of), FSM.

In these 189 countries, the period between the peak timing of the first and second waves of the SARS-CoV-2 Omicron variant ranged from 15 days in El Salvador to 351 days in Eswatini, with a median of 164 days (IQR:125.5-184.0), and generally followed a Weibull distribution (Figure 3a). In 55 (30%) countries, the period between the peak timing of the first and second waves was between 150–180 days. In only seven countries, the period between the peak timing of the first and second waves was less than 60 days. The primary reason for this could be the small peaks of the first waves in these countries. Thus, these fluctuations in data may play important roles in determining the waves of COVID-19. The peak of the second wave was higher than that of the first wave in only eight countries (i.e., Japan, Somalia, Gustemala, Federated States of Micronesia, El Salvador, Solomon Islands, Cayman Islands, and Australia); in the other 181 countries, the peak of the second wave was much lower than that of the first wave. In these 181 countries, on average, the peak of the second wave was only one-fifth that of the first wave. The relative peak of the second wave ranged from 1.0%–73.1% with a median value of 14.5% (IQR:6.9%-28.5%) and followed an exponential distribution (Figure 3b). More than three waves of the SARS-CoV-2 Omicron variant have occurred in 120 of 189 countries. The period between the peak timing of the first and third waves ranged from 30–404 days, with a median of 243 days (IQR:184.2-314.5) (Figure 3c); between the first and third waves in 69 (57.5%) countries, the period between the first and third waves ranged from 200–320 days. The peak of the third wave was higher than that of the first wave in only three countries: El Salvador, the Solomon Islands, and Japan. In the other 117 countries, the relative peak of the third wave ranged from 1%–83.6% with a median of 11.2% (IQR:4.5%-20.6%) and followed an exponential distribution (Figure 3d). Generally, the peak of the third wave was slightly lower than that of the second wave in these 117 countries.

**Figure 3.**
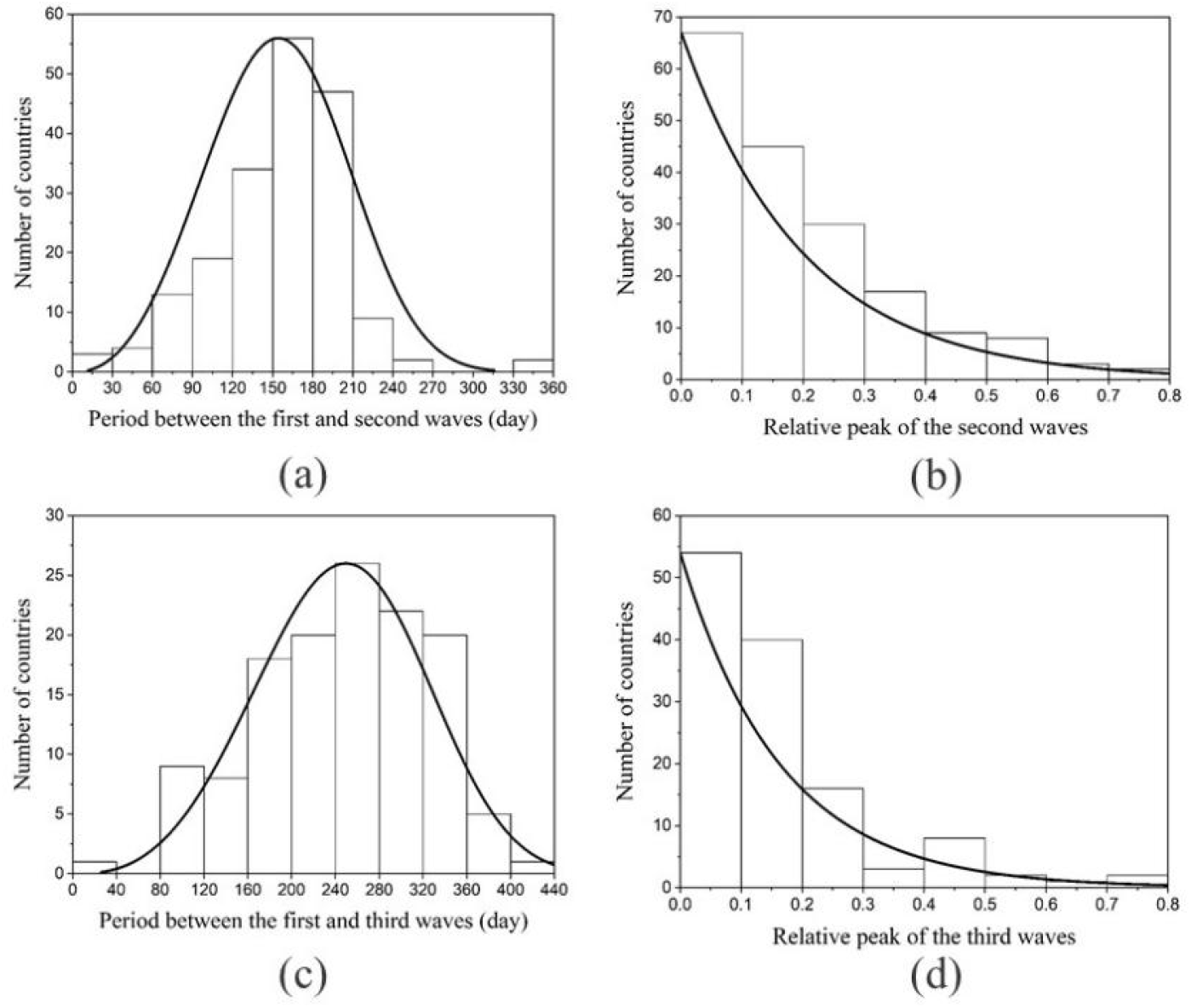
Distribution of (a) the period between the first and second waves of the SARS-CoV-2 Omicron variant in 189 countries; (b) relative peak of the second compared with the first waves in 181 countries; (c) the period between the first and third waves of the SARS-CoV-2 Omicron variant in 120 countries; and (d) relative peak of the third compared with the first waves in 117 countries.

The period between the peak timing of the first and second waves showed no statistically significant rank correlation with any of the five socioeconomic factors included in this study (Table 1). However, the relative peak of the second wave increased significantly with GDP per capita (*P*<0.001), urbanisation rate (*P*=0.003), population density (*P*=0.007), and proportion of older adults >65 years (*P*<0.001) although decreased significantly with the proportion of 0-14 teenagers (*P*<0.001) (Table 1). The main reason could be that the fast evolution of SARS-CoV-2 plays a key role in the timing of the second COVID-19 waves. If a new variant escapes the existing immune response, even a recent infection may not guarantee protection. The magnitude of the second waves could have been influenced by socioeconomic factors. Higher proportions of older adults, higher urbanisation rates, and higher population densities could accelerate the transmission of SARS-CoV-2, while a higher proportion of teenagers may suppress its spread since population susceptibility to SARS-CoV-2 increases with age.

**Table 1.** Correlation between socioeconomic factors and the timing and relative peak of the second compared with the first waves

| Socioeconomic factors | Peak timing |  | Relative peak |  |
| --- | --- | --- | --- | --- |
|  | Spearman correlation | *P* value | Spearman correlation | *P* value |
| GDP per capita* | -0.037 | 0.616 | 0.298 | <0.001 |
| Urbanization rate | 0.042 | 0.571 | 0.214 | 0.003 |
| Population density | -0.077 | 0.294 | 0.196 | 0.007 |
| Proportion of older adults >65 years | 0.127 | 0.081 | 0.307 | <0.001 |
| Proportion of <14 teenagers | -0.064 | 0.384 | -0.347 | <0.001 |

\*GDP: Gross domestic product

The SEIRS model fit the surveillance data well (Figure 4a). Moreover, was estimated at 0.62 regardless of the immunity period (6 versus 12 versus 20 months), corresponding to the reproductive number R_0_=5.44. The simulation results showed that the peak of the second wave was approximately 22% of the peak of the first wave, regardless of whether the immunity period was set at 6, 12, or 20 months. With longer immunity periods, the timing of the second wave also occurred later, and we found that with an increase in the immunity period, the timing of the onset of the second COVID-19 wave increased linearly (Figure 4b).

**Figure 4.**
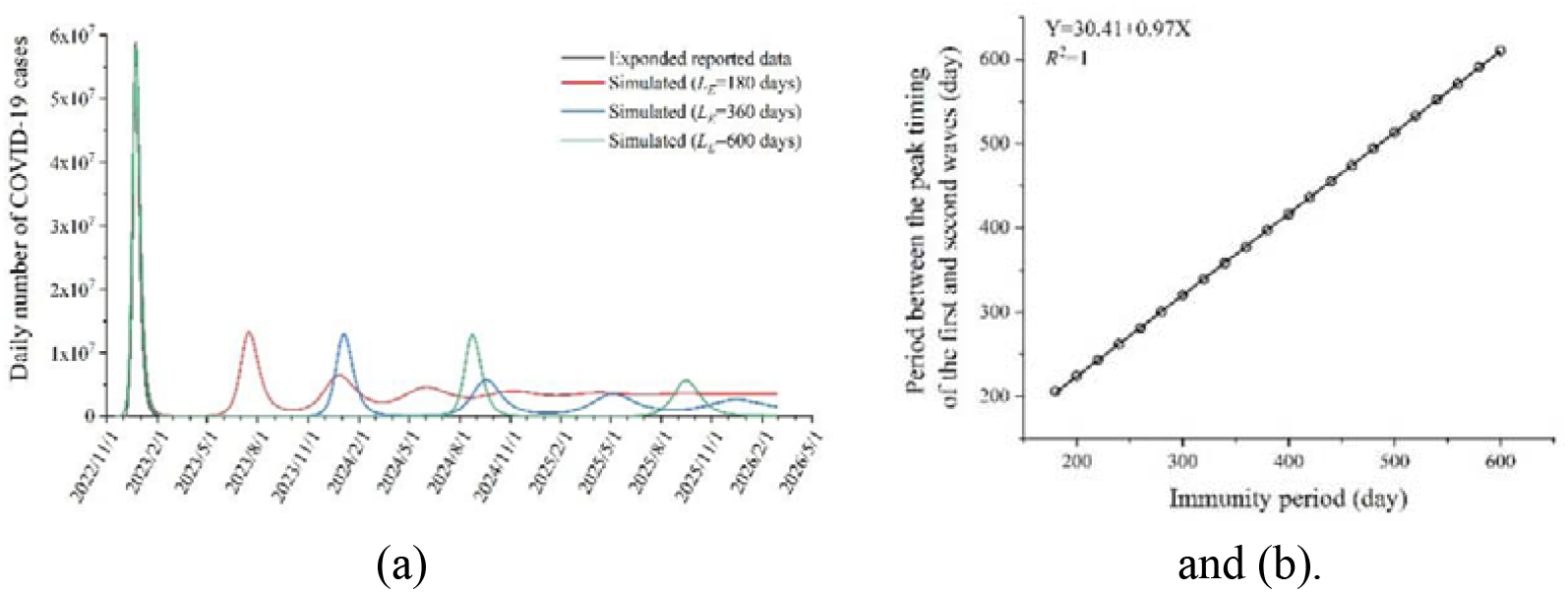
Simulated COVID-19 transmission in China with different SARS-CoV-2 immunity periods. (a) Transmission dynamics of COVID-19 and (b) timing of the onset of the second wave of COVID-19.

## Discussion

Predicting the timing and magnitude of the next COVID-19 wave is vital to better prepare for and respond to future crises. Owing to the dynamic zero-COVID policy implemented in China, the first nationwide COVID-19 wave caused by the SARS-CoV-2 Omicron variant occurred in China much later compared with other countries. Thus, we can learn from the development of COVID-19 outbreaks in other countries. In our study, we analysed the COVID-19 surveillance data from 189 countries and found that 5–7 months after the first waves of COVID-19 caused by the SARS-CoV-2 Omicron variant, the second waves occurred in most countries; however, the magnitude of the second wave was only 15% of the peak of the first wave. In this study, we also found that among 107 of 151 countries in the Northern Hemisphere, the peak timing of the second COVID-19 wave was between 1 June and 30 August 2022 which means that warm weather has limited effects on suppressing the COVID-19 epidemic caused by the SARS-CoV-2 Omicron variant [20]. This may be because, at the early stage of an emerging pathogen, a susceptible supply could limit the role of climate [21].

The median period between the first and second waves of COVID-19 was 164 days, which is slightly shorter than the reported immunity period, that is, approximately 8 months of immunity through vaccination, 12 months of immunity through infection, and longer than 12 months of immunity through both [17-19, 22]. The main reason for this could be the rapid evolution of SARS-CoV-2 [23]. If a new variant escapes the existing immune response, the recent infection may not guarantee protection [2]. We also found that socioeconomic factors affected the peaks of the second waves. With a higher GDP per capita, urbanisation rate, population density, and proportion of older adults >65 years, the peak of the second wave was larger, whereas with a higher proportion of teenagers, the peak of the second wave was smaller. This is a reasonable finding because population susceptibility to SARS-CoV-2 reportedly increases with age [9]. Thus, with the increasing numbers of older people and decreasing numbers of teenagers in the general population, SARS-CoV-2 may be transmitted quickly. A higher urbanisation rate, population density, and GDP per capita suggest better economic and societal development; thus, contact between people may also be increased, and the transmission of SARS-CoV-2 through the population may also occur quicker [9].

Finally, we must admit that more than 3 years after the COVID-19 pandemic was first identified in December 2019 [4], many unknown factors that could affect the transmission dynamics of SARS-CoV-2 remain, such as viral evaluation and environmental drivers. We found that the timing of the second wave of COVID-19 had no statistically significant rank correlation with socioeconomic factors, such as population density and urbanisation rate.

## Data Availability

All data produced are available online at:WHO Coronavirus (COVID-19) Dashboard(https://covid19.who.int), Johns Hopkins University Center for Systems Science and Engineering (https://coronavirus.jhu.edu/map.html), and Our World in Data (https://ourworldindata.org/covid-vaccinations).

https://coronavirus.jhu.edu/map.html

https://ourworldindata.org/covid-vaccinations

## References

1. 全国新型冠状病毒感染诊疗和监测数据概述(2022 年 12 月 9 日至 2023 年 1 6708 23 日) [EB/OL].2023[2023-1-25]. https://weekly.chinacdc.cn/fileCCDCW/cms/news/info/upload/ccdcw-surveillance-202212-202301-cn.pdf

2. Willyard C. How quickly does COVID immunity fade? What scientists know. Nature, 2023.

3. Cao Y, Jian F, Wang J, et al. Imprinted SARS-CoV-2 humoral immunity induces convergent Omicron RBD evolution. Nature, 2022: 1–3.

4. Bobrovitz N, Ware H, Ma X, et al. Protective effectiveness of previous SARS-CoV-2 infection and hybrid immunity against the omicron variant and severe disease: a systematic review and meta-regression. The Lancet Infectious Diseases, 2023.

5. World Bank. World Bank Open Data. 28 February, 2023. Available at: https://data.worldbank.org.cn/.

6. World Health Organization. WHO Coronavirus (COVID-19) Dashboard. 28 February, 2023. Available at: https://covid19.who.int/.

7. Dong E, Du H, Gardner L. An interactive web-based dashboard to track COVID-19 in real time. The Lancet infectious diseases, 2020, 20(5): 533–534.

8. Oxford Martin School. Our World in Data. 3 March, 2023. Available at: https://ourworldindata.org/covid-vaccinations.

9. Fan Y, Li X, Zhang L, et al. SARS-CoV-2 Omicron variant: recent progress and future perspectives. Signal transduction and targeted therapy, 2022, 7(1): 141.

10. Lei H, Yang L, Wang G, et al. Transmission Patterns of Seasonal Influenza in China between 2010 and 2018. Viruses, 2022, 14(9): 2063.

11. Yang W, Cowling B J, Lau E H Y, et al. Forecasting influenza epidemics in Hong Kong. PLoS computational biology, 2015, 11(7): e1004383.

12. China Statistical Yearbook. 28 February, 2023. Available at: http://www.stats.gov.cn/tjsj/ndsj/2022/indexch.htm.

13. Xin, H. et al. Transmission dynamics of SARS-CoV-2 Omicron variant infections in Hangzhou, Zhejiang, China, January-February 2022. International Journal of Infectious Diseases 126, 132–135 (2023).

14. Zheng, Y. & Wang, Y. Transmission Characteristics and Predictive Model for Recent Epidemic Waves of COVID-19 Associated With OMICRON Variant in Major Cities in China. Int J Public Health 67, 1605177 (2022).

15. Huang J, Zhao S, Chong K C, et al. Infection rate in Guangzhou after easing the zero-COVID policy: seroprevalence results to ORF8 antigen. The Lancet Infectious Diseases, 2023.

16. Michlmayr D, Hansen C H, Gubbels S M, et al. Observed protection against SARS-CoV-2 reinfection following a primary infection: A Danish cohort study among unvaccinated using two years of nationwide PCR-test data. The Lancet Regional Health-Europe, 2022, 20: 100452.

17. Marking U, Bladh O, Havervall S, et al. 7-month duration of SARS-CoV-2 mucosal immunoglobulin-A responses and protection. The Lancet Infectious Diseases, 2023.

18. Chemaitelly H, Tang P, Coyle P, et al. Protection against Reinfection with the Omicron BA. 2.75 Subvariant. New England Journal of Medicine, 2023.

19. Nordström P, Ballin M, Nordström A. Risk of SARS-CoV-2 reinfection and COVID-19 hospitalisation in individuals with natural and hybrid immunity: a retrospective, total population cohort study in Sweden. The Lancet Infectious Diseases, 2022, 22(6): 781–790.

20. Bashir MF, Ma B, Komal B, et al. Correlation between climate indicators and COVID-19 pandemic in New York, USA. Science of the Total Environment, 2020, 728: 138835.

21. Baker R E, Yang W, Vecchi G A, et al. Susceptible supply limits the role of climate in the early SARS-CoV-2 pandemic. Science, 2020, 369(6501): 315–319.

22. Hall V, Foulkes S, Insalata F, et al. Protection against SARS-CoV-2 after Covid-19 vaccination and previous infection. New England Journal of Medicine, 2022, 386(13): 1207–1220.

23. Callaway E. Fast-evolving COVID variants complicate vaccine updates. Nature, 2022, 607(7917): 18–19.

